# *KinformR*: novel pedigree and candidate variant scoring methods for family-based genetic studies

**DOI:** 10.1101/2025.10.06.25337426

**Authors:** Cameron M Nugent, Margaret MacMillan, Tom Barber, Bari J Ballew

## Abstract

**Summary:** Family-based genetic studies are employed to identify rare variants underlying heritable diseases. In practice, these studies present challenges in both participant enrollment and variant prioritization. Family-based studies often include many pedigrees that can be difficult to prioritize, and within families, not all family members are equally important as subjects for variant analysis. Understanding the extent of penetrance and the proportion of the genome with identity-by-descent (IBD) can show the relative value of different pedigrees, and of different subjects within a pedigree. Additionally, analyzing multiple pedigrees for association between genotypes and disease status can reveal several candidate variants whose relative merits can be difficult to weigh against one another. Incorporating the degree of relatedness of individuals with their genotypes and disease status can provide an additional metric when evaluating variants within a family, accounting for both incomplete penetrance and segregation with the phenotype.

*KinformR* is an R package that provides relationship-informed scoring metrics to aid in the design and interpretation of family-based genomic analyses. The package provides two main scoring functions meant to aid in: 1) pre-analysis evaluation of pedigrees to direct recruitment and sampling efforts by quantifying their potential relative detection power, and 2) post-sequencing prioritization of candidate variants. By facilitating comparison of the relative detection power of families in a study and the value of candidate variants’ association with disease status, the package provides relationship-informed complements to existing analysis tools that focus on the genomic consequences of variants.

To demonstrate the utility of these new methods, several example families have been analyzed using *KinformR*. The *KinformR* scoring metrics offer novel statistical methods for enhancing efficiency and accuracy of rare variant association studies and elucidating the genetic architecture of diseases in family-based data.

## Introduction

Family-based sequencing studies are effective means of identifying the genetic basis of heritable diseases [1,2]. Families containing multiple affected individuals increase detection power relative to population-based studies through the ability to discern rare variation segregating with disease phenotypes [3,4]. Additionally, family-based designs contain both within- and between-family information [5] and facilitate incorporation of genotypes and phenotypes from large pedigrees [5]. Although family-based approaches control for common population-based analysis issues such as multiple testing [5] and population stratification [6], family studies are not without confounds, including technical artifacts, mis-reported relationships, chance associations, and incomplete penetrance, all of which can make isolating the true genetic basis of a disease challenging [7,8].

Whole genome sequencing (WGS) allows for detailed, direct empirical assessment of the genetic makeup of families [9]. The scale at which the genome can be queried allows inheritance of rare variants to be observed across related individuals [9,10] and association of genetic variants with disease status has enabled discovery of numerous novel disease-associated genes through family-based studies [11–13]. However, WGS’s increased resolution is accompanied by elevated rates of false genotype-phenotype associations [14], which can make it difficult to identify genetic causes of disease, even with proper quality control measures [11–13,15-17].

The fundamental unit of analysis in family-based studies is the genetically related family structure, as represented on a pedigree; based on the relationships of individuals, it is possible to measure the prevalence, transmission, and penetrance of a disease and associate the phenotype to underlying genetic variants. Penetrance is a measure of the probability that an individual possessing a disease-causing variant will manifest the corresponding disease phenotype [18,19]. Incomplete penetrance provides a confound in family-based genetic studies, leading to a decoupling of phenotype-genotype associations, with unaffected individuals potentially possessing a variant [18,19]. Pedigree-based estimation of autosomal dominant penetrance was proposed by Rogatko et al. [20] and refined by Horimoto et al. [7]. Understanding the extent of penetrance is important information that can be considered when searching for the genetic underpinnings of disease; incomplete penetrance can be observed in both autosomal dominant and recessive conditions [21], though the present study focuses on the former. The proportion of the genome shared between individuals due to being inherited from a common ancestor is quantified through the metric of identity-by-descent (IBD) [22]. At the population level, measuring IBD has given insights into demographic history, population structure, and natural selection [23,24]. On more proximal scales, estimating IBD across pedigrees can show the amount of the genome shared across individuals through chance inheritance and give an indication of the false positive rate when associating genotypes and disease phenotypes. Estimation of penetrance and IBD can show the power of pedigrees for disease-associated rare variant discovery.

Association between genotypes and disease status in family-based studies often reveals multiple plausible candidate variants. Understanding the biological consequences of rare variants via annotation tools like the Ensembl Variant Effect Predictor (VEP) [25] is one means of evaluating suitability of rare variant candidates, as is assessing *in silico* predictions of pathogenicity with tools like SIFT [26] or CADD [27]. These tools provide context-informed evidence of biological consequences of variants, although variants can be overlooked if their effects are subtle, or they fall in poorly described genomic regions [11–13,27,28]. The degree of relatedness of individuals and segregation of variants provides an additional line of evidence to consider when evaluating rare variants in familial disease studies that is not considered by consequence predictors. For example, in family-based case-control studies, use of sibling controls leads to estimates of genetic relative risk that are approximately half as efficient as those obtained with the use of population controls, while relative efficiency for cousin controls is approximately 90% [29,30]. When working with large, complex pedigrees, relationship-informed assessment of variant segregation can complement consequence-based scoring tools and provide an additional stream of empirical evidence, thereby allowing for a more holistic view of potential variant-disease associations.

*KinformR* is an R package meant to aid in comparative evaluation of families and candidate variants in rare-variant association studies, by providing relationship-informed pedigree and variant scoring. The package can be used for two methodologically overlapping but distinct purposes: 1) prior to any genetic/genomic analysis, evaluation of relative discovery power of pedigrees can direct recruitment efforts by showing which families have the best potential for discovery and which unsampled individuals would be the most meaningful additions to a study, and 2) after sequencing and analysis, scoring variants based on association with disease status and familial relationships of individuals to aid in variant prioritization. Both functions are based on evaluating IBD between related individuals to generate scores, and penalizing for incomplete penetrance. The package can improve family-based studies in early stages by directing resource allocation and sampling effort. Following analysis, scoring of variants can aid in the empirical description and comparison of candidate variants, which can help inform decisions about the most promising variants to dedicate time and resources towards.

## Pedigree scoring

### Simplifying assumptions

The initial release of *KinformR* incorporates several simplifying assumptions to calculate penetrance and IBD in families, specifically: i) the trait under study is autosomal dominant, ii) all individuals have unambiguous disease status (i.e. affected or unaffected), iii) in cases where carrier status is unknown (i.e. an unaffected individual can either be a non-obligate carrier or a non-carrier), if there are three or more sequential generations of such individuals, they shall be considered unaffected, and iv) underlying true penetrance is constant (e.g. unaffected by other factors such as environment). Individuals less than the age of onset of the condition under study are excluded from analysis, as they cannot be assigned unambiguous disease status (see (ii)). We recommend excluding large branches of unaffected individuals for simplicity; including pedigree structures with three or more sequential generations of unaffected individuals (see (iii)). This slightly biases the result towards a higher estimate of penetrance.

### Encoding of the pedigree

A frequentist approach is utilized to calculate penetrance via a likelihood function for each pedigree [31,32]). The calculation relies on encoding individuals into the following categories:

*A* : Affected individuals

*B* : Obligate carriers

*C* : Children of affected individuals or carriers, with no children of their own

*D* : Trees of unaffected individuals - specifically, two sequential generations

(i.e. parent and offspring; larger trees of unaffected individuals are omitted.)

To encode **Figure 1A**, (modified version of pedigree from Otto & Harimato 2012), counts of individuals are made for categories A, B, and C. There are four individuals in category A (III-C, III-G, III-H, and IV-A), four individuals in B (I-A, II-B, II-C, III-F = 4), and three individuals in C (III-D, III-E, III-I). Category D is represented by two numbers: the number of offspring in a tree of unaffecteds, d, and the number of trees that have that same number of offspring in the pedigree, n. The pedigree in **Figure 1A** has one tree with two offspring (d=2, n=1).

**Figure 1.**
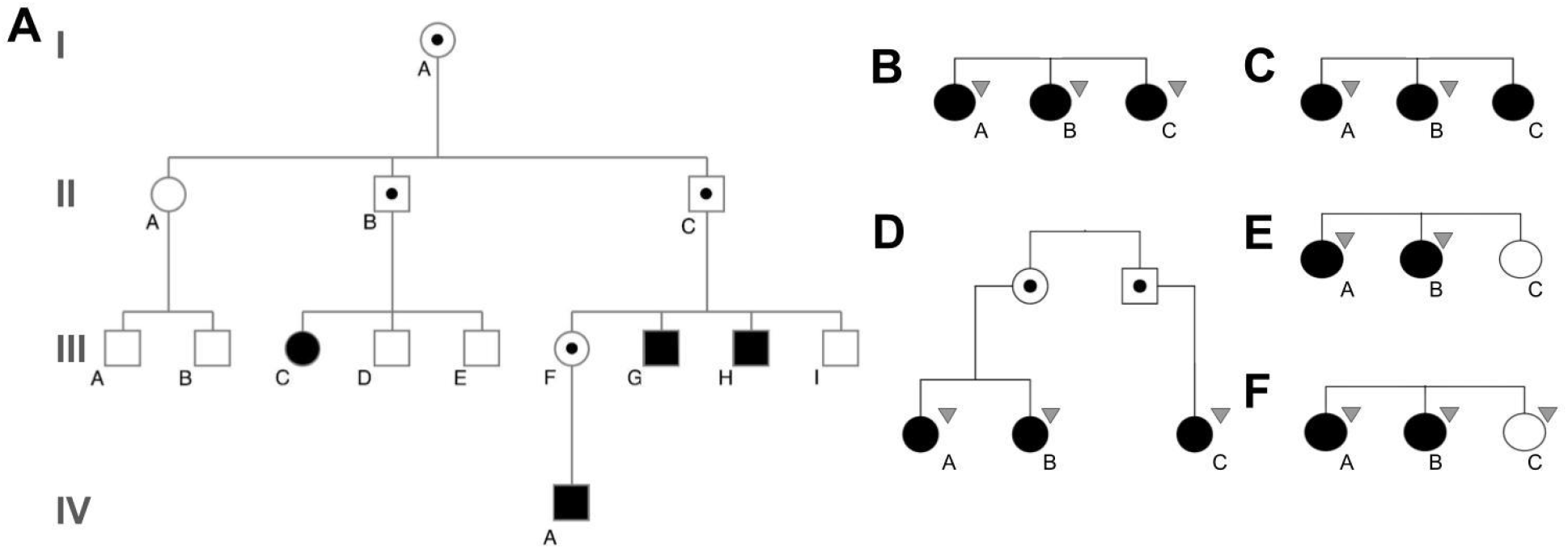
For all diagrams, dark filled symbols represent affected individuals, light coloured symbols represent unaffected individuals, and dark dots in the centre of symbols represent obligate carriers. Grey triangles represent individuals carrying a disease-associated variant. **A**. Example pedigree adapted from Otto & Harimato (2012). Individual IDs are indicated by the roman numerals indicating the generation and the letters underneath the individuals. **B**. Three affected siblings, all of whom have a variant. **C**. Three affected siblings, two of whom have a variant. **D**. Three affected individuals, a sib pair and cousin, sharing a common variant. **E**. A three sibling family with one unaffected sibling not carrying the candidate variant. **F**. A three sibling family with all individuals possessing the candidate variant.

### Combining IBD and penetrance

The combined scoring metric is based on penetrance and IBD (for derivation of the metric and its constituents, see Supplementary File S2) associated with a pedigree that can be used to quantify both the prospective (theoretical maximum) and realized (current sample set) value of a family for association analysis. This metric allows comparison of the relative “power” of families in a study; the score goes up as the search space is narrowed, in proportion to IBD, and the score is penalized for incomplete penetrance, which increases the search space due to uncertainty.

## Variant scoring

Relatedness of individuals can also be used to score the value of a candidate variant’s association with disease. A candidate is strongest when shared by an affected individual and distantly-related affected relatives, and not shared by closely related unaffected individuals. More distant relatives have smaller proportions of their genome IBD, so there is a lower chance of false positives (shared variants not associated with disease). Conversely, closely related affected and unaffected individuals provide evidence that the disease is not associated with the large IBD portion of their genomes.

Variant scoring determines a candidate variant’s merit by considering: phenotype status (affected/unaffected), candidate variant genotypes, and familial relationships.

At its most basic, scoring is conducted from a reference individual’s perspective. In **Figure 1B**., several siblings are affected and possess a variant of interest. Using individual *A* as the reference individual, the variant is scored based on shared genotype and affected status.

Affected scores are calculated as the reciprocal of the coefficient of relatedness. Individuals *A* and *B* are 1st degree relatives (coefficient of relatedness (*r*) of 0.5) obtaining a score of:

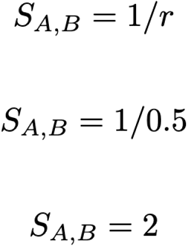

The score for *A* and *C* is the same. Lastly, *A* is scored relative to themselves (coefficient of relatedness of 1) giving the variant a total score of 5 points:

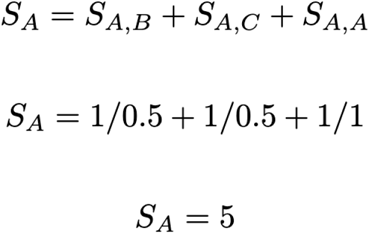

**Figure 1C**. shows a weaker candidate, shared by *A* and *B*, but not *C*. In this case, *evidence for* and *evidence against* are combined, quantifying this candidate as weaker.

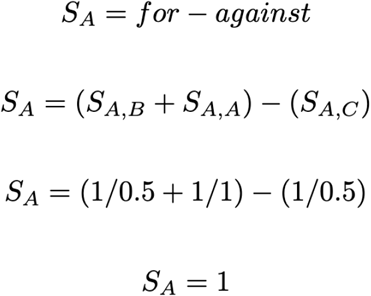

For the previous three sibling example, all perspectives are equal. However, when pedigrees are more complex, scores for candidates change depending on the reference individual. In **Figure 1D**., scoring using *A* as the reference describes *B* as a first degree relative and *C* as a 3rd degree relative (coefficient of relatedness of 0.125), yielding the score:

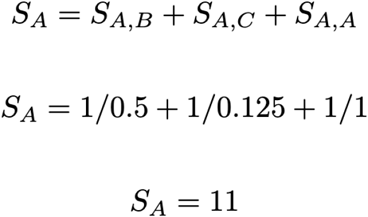

Whereas the perspective of *C* yields a higher score because both *A* and *B* are 3rd degree relatives.

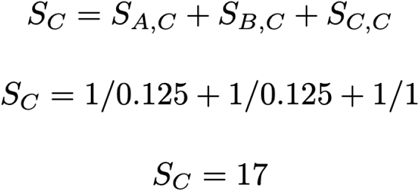

Therefore, variant scoring is calculated using each affected individual as the reference and averaging perspectives to obtain a final score. For **Figure 1D**., this is:

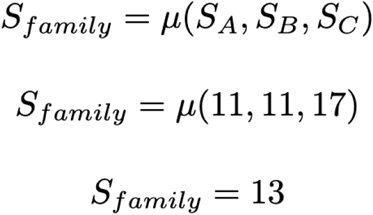

### Unaffected individuals

For scoring unaffected individuals, directionality of evidence is inverted and we consider the absence of candidates in individuals who are not expected to possess a candidate variant. Evidence for a candidate comes from eliminating alternative hypotheses by showing IBD variants are likely not to be causative. For an affected reference and an unaffected sibling, half their genomes are IBD, giving evidence against any of these shared variants being causative. As the coefficient of relatedness decreases, smaller fractions of the genome are IBD and therefore, scores decay the more distantly related an unaffected individual is from the reference based on the formula:

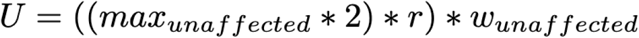

The default `unaffected.max` value is 8, which defines the maximal score that closely related unaffected individuals can provide for a variant. Evidence for unaffected individuals is confounded by incomplete penetrance. A completely penetrant variant would not be present in unaffected individuals in a pedigree, but unaffecteds may possess a variant if penetrance is lower. To account for this, unaffected evidence is by default given half the weight of affected evidence (unaffected_weight = 0.5). The weighting can be customized based on penetrance estimates or removed entirely to consider only affecteds.

Scoring **Figure 1E**. from perspective of *A*, the unaffected sibling is scored as:

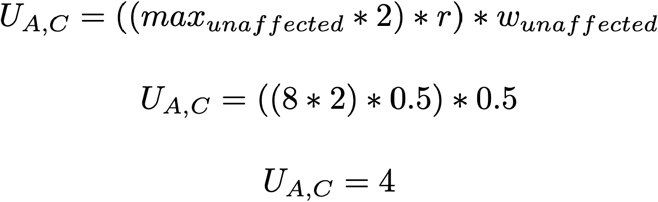

Affected and unaffected points are summed.

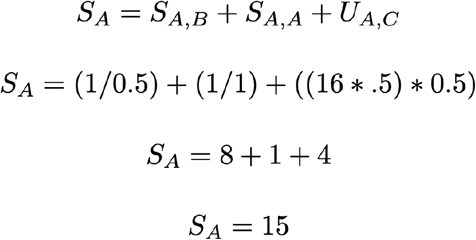

If the unaffected individual has the variant (**Figure 1F**.), the score for the *A,C* relationship would be *evidence against* the candidate.

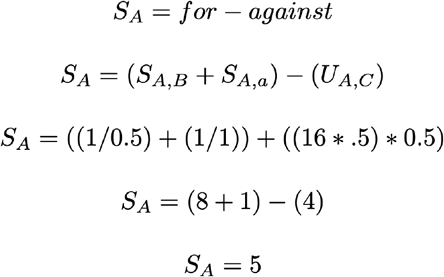

## Discussion

The relationship-informed pedigree and variant scoring methods of *KinformR* expand the range of evaluation tools available for family-based genetic studies. The accompanying case study (Supplemental Files S2 and S3) demonstrates practical utilization of *KinformR*, through examination of four pedigrees and accompanying candidate variants.

During study design, careful consideration of information contained within pedigrees can reveal the prevalence and penetrance of a disease as well as the putative detection power of a family. Sample collection, DNA extraction, sequencing, and bioinformatics analysis are costly in terms of time, funds, and person hours. Therefore, conducting these steps in a deliberate manner increases the chance of uncovering true variant-disease relationships. *KinformR*’s family scoring metrics provide means for increasing efficiencies associated with design and management of family-based association analyses. Specifically, prospective and realized pedigree scores are of value for: i) determining which families to focus recruitment efforts on, ii) which non-sampled individuals would be the most informative additions to the score, iii) determining the value of sequencing different individuals.

When multiple, potentially private [33] candidate variants are identified across different families in a study, it can be challenging to compare their relative value and make decisions about allocating time and resources for follow-up analysis. Scoring methods such as SIFT [26] and CADD [27] are based on the biological implications of variants. The *KinformR* variant scoring method is not a replacement for these tools, rather a compliment to them that uses family structure and relationship information to provide an additional line of evidence to consider when evaluating rare variants in family-based studies.

The package *KinformR* serves as empirical bookends, increasing the efficiency of resource allocation, sampling effort, and iterative analysis of family-based variant data. By facilitating informed decisions about the most promising families and variants to dedicate time and resources towards, the *KinformR* package and IBD-based scoring methods therein improve the efficiency and chance of successfully identifying the genetic basis of rare diseases.

## Availability and Implementation

R package and vignettes are available via GitHub: https://github.com/SequenceBio/KinformR

## Contact

For software related inquiries: https://github.com/SequenceBio/KinformR

Additional inquiries can be directed to

## Supplementary Information

Supplementary File S1 - Derivation of penetrance estimation algorithm

Supplementary File S2 - Case study example use of KinformR R package

Supplementary File S3 - R code and test data accompanying the case study example use

## Data Availability

All data produced in the present work are contained in the manuscript.

https://github.com/SequenceBio/KinformR

