## Supplementary_File_S1 for "*KinformR*: novel pedigree and candidate variant scoring methods for family-based genetic studies"

**Supplementary File S1.** Derivation of penetrance estimation algorithm.

If *K* is the penetrance rate, the probability of a condition affecting from each group is:

a. For affecteds, K multiplied by the probability of inheriting the variant from one parent, i.e.:

$$\frac{1}{2} * K = \frac{K}{2}$$

b. For obligate carriers, the probability is the inverse proportion of the penetrance rate, i.e.:

$$\frac{1}{2} * (1 - K) = \frac{(1-K)}{2}$$

c. The pedigree progenitor is a special case, calculated without considering inheritance from previous generations, i.e.:

$$1 - K$$

d. For unaffected offspring, it is unknown if they are carriers. Either they inherited the non-disease allele from the carrier parent `(1/2)`, or they inherited the disease-associated allele but did not develop the disease `(1/2 * (1-K))`, i.e.:

$$\begin{aligned}\left(\frac{1}{2} * (1-K)\right) &= \frac{1}{2} + \frac{1}{2} * (1-K) \\ &= \frac{(2-K)}{2}\end{aligned}$$

The likelihood function is the product of the probabilities for these groups:

$$L = \left[\frac{K}{2}\right]^a * \left[\frac{(1-K)}{2}\right]^b * \left[\frac{(2-K)}{2}\right]^c * \left[\frac{(2^n + (1-K) * (2-K)^n)}{2^{n+1}}\right]^d$$

Where a, b, and c are the numbers of individuals in these categories, for d, the formula accounts for multiple branches and variable numbers of second generation individuals on these branches.

$$
\begin{aligned}
L &= \left[\frac{K}{2}\right]^a * \left[\frac{(1-K)}{2}\right]^b * \left[\frac{(2-K)}{2}\right]^c * \left[\frac{(2^n + (1-K)*(2-K)^n)}{2^{n+1}}\right]^d \\
LL &= \log(L) \\
&= \log\left(\left[\frac{K}{2}\right]^a\right) + \log\left(\left[\frac{(1-K)}{2}\right]^b\right) + \log\left(\left[\frac{(2-K)}{2}\right]^c\right) + \log\left(\left[\frac{(2^n + (1-K)*(2-K)^n)}{2^{n+1}}\right]^d\right) \\
&= a*\log\left(\frac{K}{2}\right) + b*\log\left(\frac{(1-K)}{2}\right) + c*\log\left(\frac{(2-K)}{2}\right) + d*\log\left(\frac{(2^n + (1-K)*(2-K)^n)}{2^{n+1}}\right)
\end{aligned}
$$

The equation is simplified by taking the log and removing constants with respect to K (solving for maximum log is equivalent to solving for the maximum, if maximum is non-zero). Thus, the maximum likelihood estimate of penetrance (`K`) for the family is obtained:

$$
\begin{aligned}
LL \sim\ & a*\log(K) + b*\log(1-K) + c*\log(2-K) \\
& + d*\log(2^n + (1-K)*(2-K)^n) - d*(n+1)*\log(2)
\end{aligned}
$$

The term d must be calculated for each tree of unaffecteds and summed:

$$
\begin{aligned}
LL \sim\ & a*\log(K) + b*\log(1-K) + c*\log(2-K) \\
& + \sum(d*\log(2^n + (1-K)*(2-K)^n) - d*(n+1)*\log(2))
\end{aligned}
$$

*Estimating IBD*

Theoretical IBD is a function of Wright's coefficient of relatedness, calculated by counting the number of non-overlapping paths between pedigree members (either all pedigree members for a maximum potential IBD, or only sampled pedigree members for actual IBD), which is `n` in:

$$(1/2)^n$$

This method is not constrained to estimating IBD from a single perspective (i.e. that of the proband) but rather considers all relationships in the pedigree.

When calculating relatedness, certain relationships are penalized based on the penetrance value, K. Category C (children of obligate carriers or affecteds) have their relatedness contribution multiplied by K. For category D (trees of unaffecteds), if the parent is sampled, they are treated the same as a category C individual. If the parent is not sampled by but one or more children are, we penalize the child's relatedness contribution by K^2 (accounting for non-penetrance).

The formula for estimation of realized (sampled) IBD is:

$$x = \sum (d * (1 - (2^n - 1)/2^{n+1})) * 2^K$$

$$\hat{\pi} = (a + b + c * 2^K + x) - 1$$

the formula i for theoretical maximal IBD is:

$$x = \sum (d) * 2^K$$

$$\hat{\pi} = (a + b + c * 2^K + x) - 1$$

The final combined scoring metric based on penetrance and IBD is calculated as:

$$\log(2^{\hat{\pi}})$$
