## Supplementary_File_S2 for "*KinformR*: novel pedigree and candidate variant scoring methods for family-based genetic studies"

**Supplementary File S2. Case Study Example Use**

***Pedigree scoring***

Here we present a small-scale example of the use of the pedigree and variant scoring functions for evaluating a set of four families. See Supplementary File S3 for associated R code and input data files. For additional examples of use of the *KinformR* package, see the package's vignettes (https://github.com/SequenceBio/KinformR).

Within **Figure S1,** pedigrees for the four families of the example study are presented. To assess the levels of penetrance and IBD in the families, the data are first encoded into the scoring inputs (**Table S1.**) The encoding includes two sets of values: the max, where all individuals, sampled or not, are included in the encoding, and the sampled, where only those individuals for which samples have been collected are encoded.

Processing the inputs with *KinformR* provides the outputs shown in **Table S2.** From this information, we can see that with the samples collected, Fam4 has the highest realized variant detection power, with a score of 2.75. However, were sampling efforts to be expanded, Fam1 has the highest potential detection power, with a max score for the family of 6.76 if data were collected for every individual in the pedigree.

***Variant scoring***

To demonstrate the variant scoring methods, Figure S1. also presents the genotypes for candidate variants in the sampled individuals. The disease status, genotypes, and relationships of the individuals can then be encoded for analysis with *KinformR*. Outputs of variant scoring for the variants with default parameters is shown in **Table S3.**

The scores reveal the strongest intra-family evidence for a variant to be for the variant in Fam2, with a cumulative score of 7.33. This is due to the variant being shared between two affected cousins and not present in one of their unaffected offspring. However the absence of the variant in the third affected cousin provides evidence against the variant, which should also be considered in prioritization of followup. Overall Fam2 has the potential for detecting the strongest candidate variants, as the theoretical max score for a variant in this family is 18.67 (see accompanying R script for the theoretical), which would be achieved if the variant were also present in the third affected cousin.

Within **Table S3.** the variant column shows that the same candidate variant (v3) is observed in both Fam3 and Fam4. These scores can be summed together, to obtain a cross family score for the variant of 3 + 6.5 = 9.5. This shows that this candidate variant is in fact stronger than the candidate exclusive Fam2 when evidence is pooled across families. Although the affected individuals in the two pedigrees are more closely related, and provide weaker evidence on their own, we can see that when the evidence is pooled there is in fact stronger support for this candidate variant's association with disease compared to the variants exclusive to the first two families. The absence of this variant in individual Fam2-1003 suggests that it may be a false positive, as the variant is not present within the third branch of the family that has been sampled.

A

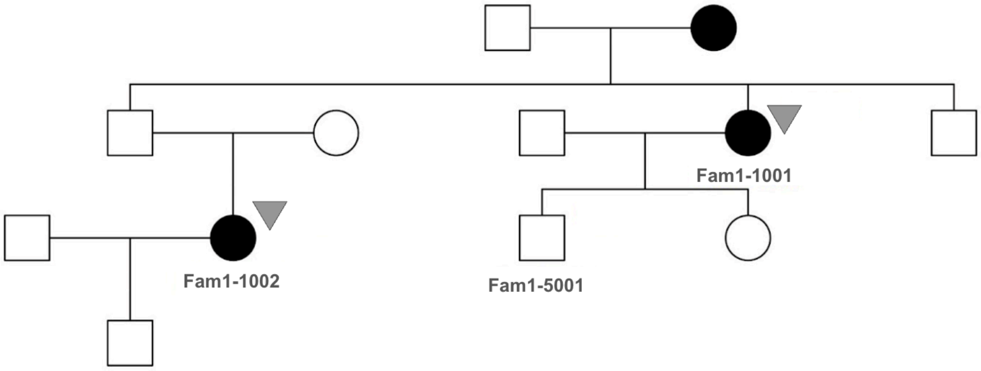

Fam1-1001
Fam1-1002
Fam1-5001


B

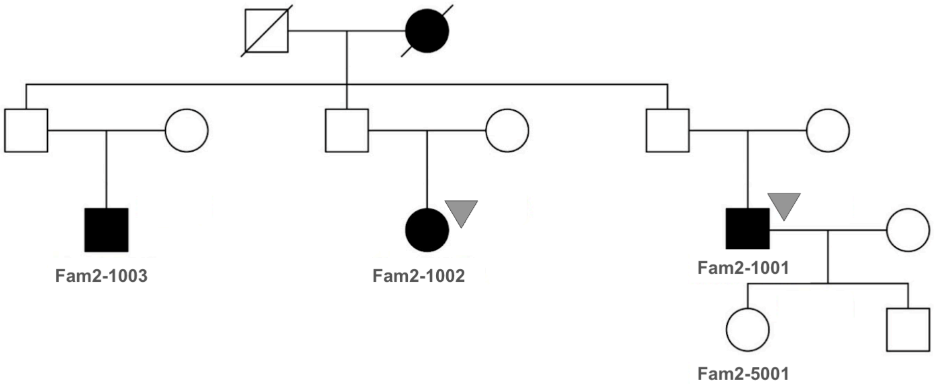

Fam2-1003
Fam2-1002
Fam2-1001
Fam2-5001

C

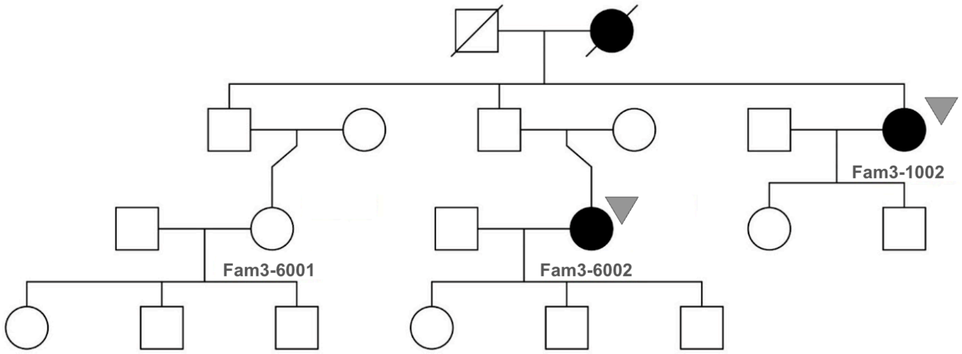


D

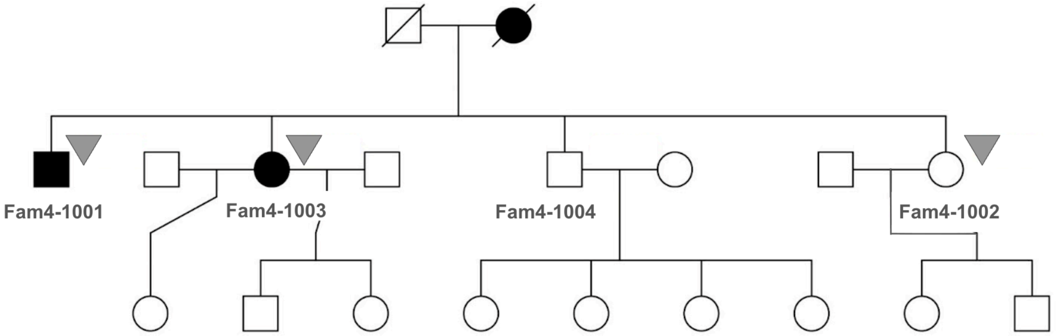


**Figure S1.** Four families of the case study. Individuals with an ID are those for whom samples have been collected. Grey triangles to the top right of sampled individuals denotes those possessing the given candidate variant in the family.

**Table S1.** Encoding of study families for pedigree scoring.

| family | a | | b | | c | | d | | n | |
|---|---|---|---|---|---|---|---|---|---|---|
| | max | sampled | max | sampled | max | sampled | max | sampled | max | sampled |
| Fam1 | 2 | 2 | 1 | 1 | 5 | 0 | 1 | 1 | 1 | 1 |
| Fam2 | 3 | 2 | 1 | 0 | 4 | 1 | 0 | 0 | 0 | 0 |
| Fam3 | 3 | 3 | 3 | 0 | 2 | 1 | 0 | 0 | 0 | 0 |
| Fam4 | 2 | 2 | 0 | 0 | 3 | 0 | 2 | 2 | 0 | 0 |

**Table S2.** Scoring outputs for the four families of the case study

| Family | Penetrance | Max pi-hat | Max Score | Current Pi-hat | Current score | Realized percent of max score |
|---|---|---|---|---|---|---|
| Fam1 | 0.37 | 9.76 | 6.76 | 2.97 | 2.06 | 30.44 |
| Fam2 | 0.58 | 8.97 | 6.22 | 2.49 | 1.73 | 27.79 |
| Fam3 | 0.45 | 7.73 | 5.36 | 3.36 | 2.33 | 43.53 |
| Fam4 | 0.57 | 8.43 | 5.84 | 3.97 | 2.75 | 47.12 |

**Table S3.** Candidate variant scores in the Case Study families

| Family | Variant | Total score | Score for | Score against |
|---|---|---|---|---|
| Fam1 | v1 | 6 | 6 | 0 |
| Fam2 | v2 | 7.33 | 13 | 5.67 |
| Fam3 | v3 | 6.5 | 6.5 | 0 |
| Fam4 | v3 | 3 | 7 | 4 |
