## Supplementary_File_S3 for "*KinformR*: novel pedigree and candidate variant scoring methods for family-based genetic studies": supp_example.R

```
###
# Load the package
library(KinformR)

###
#Ensure your working directory is set to the supplementary_s2 folder

###
# Load the encoded pedigree data file
example.pedigree <- read.pedigree("./pedigree_data.tsv")

###
# Score the pedigrees
example.pedigree.scores <- score.pedigree(example.pedigree)

###
# Show the results, can write to a tsv
example.pedigree.scores

###
# Parallelized scoring of the variants from the four families
# see: vignette('KinformR-variant_scoring') for detailed examples of singular execution

# Load the ID, disease, and genotype files
tsv.names <- c("Fam1.tsv", "Fam2.tsv", "Fam3.tsv", "Fam4.tsv")
indv.df <- lapply(tsv.names, read.indiv)

# load the relationship matrix files
mat.fnames <- c("Fam1.mat", "Fam2.mat", "Fam3.mat", "Fam4.mat")
mat.df <- lapply(mat.fnames, read.relation.mat)

# Determine category from disease status and genotype
status.df <-  lapply(indv.df, score.variant.status)

#score the variants
bulk.scores <- mapply(score.fam, mat.df, status.df)

# convert the scores to a dataframe
bulk.df <- data.frame(bulk.scores)
names(bulk.df) <-c("Fam1", "Fam2", "Fam3", "Fam4")

# re-score, using the theoretical.max param
# shows the max a perfect variant in each family could achieve.
theory.status.df <-  lapply(indv.df, score.variant.status, theoretical.max = TRUE)
theory.bulk.scores <- mapply(score.fam, mat.df, theory.status.df)
theory.bulk.scores.df <- data.frame(theory.bulk.scores)
names(theory.bulk.scores.df) <-c("Fam1", "Fam2", "Fam3", "Fam4")
theory.bulk.scores.df
#transpose so families are rows
t(theory.bulk.scores.df)
```
